# Risk and Uncertainty Communication in Deployed AI-based Clinical Decision Support Systems: A Scoping Review

**DOI:** 10.1101/2024.12.06.24318489

**Authors:** Nicholas Gray, Helen Page, Iain Buchan, Dan W. Joyce

## Abstract

Clinical decision support systems (CDSS) employing data-driven technology such as artificial intelligence, machine- and statistical-learning are increasingly deployed in healthcare settings. These systems often provide clinicians with diagnostic, prognostic, or risk scores modelled from curated patient-level data and frequently involve iterative and non-deterministic optimisation of flexible, parameterised models. All of these data and algorithms have uncertainties associated with them that should be taken into account when used to support clinical decisions at the patient level. This scoping review aims to describe the literature on how deployed data-driven CDSSs present information about uncertainty to their intended users. We describe common clinical applications of CDSSs, characterise the decisions that are being supported, and examine how the CDSS provides outputs to end users, including uncertainty at the individual patient level, as well as indirect measures such as CDSS performance against a reference standard. We conclude with a discussion and recommendations on how CDSS development can be improved.

## 1 Introduction

Clinical decision support systems (CDSSs) are important tools in modern healthcare used to augment clinicians’ decision-making processes [154]. With the rapid advance in data-driven technologies – notably, artificial intelligence (AI) and machine learning (ML) alongside more traditional statistical learning – their incorporation into CDSSs is frequently described as having the potential to revolutionise healthcare by augmenting human decision-making to improve diagnostic accuracy and personalised treatment.

The outputs of these systems, often probabilistic in nature, encapsulate various forms of uncertainty [80]; but there is ambiguity in how risk data and different model predictions are presented and this can be confusing for the end user. For example, QRISK3^1^ [70] calculates the risk of a patient having a heart attack or stroke over the next 10 years. QRISK3 uses 21 predictors and outputs the resulting risk of an event as a percentage and with a natural frequency style expression^2^. The same natural frequency representation is presented visually as an icon array showing the risk of an event alongside a comparison to a “healthy” individual (of the same age, ethnicity and sex) delivered as a numerical relative risk or risk ratio (i.e. the ratio of the patient and the “healthy” person’s QRISK score). These methods of communicating risk need to be clear to the end user of the tool as different expressions of uncertainty may alter comprehension, immediate decisions, and consequent actions.

Incomplete medical knowledge, which affects the labelling of data [32] used to develop CDSS algorithms, or the complexity of interacting and comorbid diagnoses/conditions [72], can also introduce uncertainty into an algorithm. Accurately representing and effectively communicating this uncertainty in CDSS algorithms is crucial, as it should influence clinical decision-making and may affect clinician and patient trust in AI recommendations [19, 62, 133]. There are numerous approaches that could be used to express the uncertainty of a clinical outcome all of which have potential benefits and drawbacks, although there is little consensus as to what the best approach would be [147].

The increasing accessibility of programming tools that can embed prediction models in clinical workflows has made creating data-driven clinical decision support systems relatively straightforward. The first step is to pick a clinical decision that the tool will support and then, locating and curating the relevant data and choosing a suitable algorithm to be used to fit the model to the data. Once this has been completed the model needs to be evaluated and, hopefully, it can be concluded that the clinical decision support tool is appropriate for use. The final step is the deployment of the tool, requiring constructing a user interface so that the tool can be used fluently in clinical practice. Then the model needs to be maintained as many lose performance with drifts in population health and data [69]. This review focuses on deployed systems as it is assumed that the creators have had to at least consider the communication of the tool’s outputs to the end-user.

This scoping review aims to explore the representation of uncertainty in *deployed* data-driven and AI-based clinical decision support systems. By systematically examining the existing literature, we seek to identify current practices, highlight challenges, and propose directions for future research.

### 1.1 Defining deployed AI based Clinical decision support systems

For the purposes of this scoping review, we will make use of the following three definitions.

**Definition 0.** A clinical decision support system is based on an algorithm designed to aid a medical decision or augment the decision making process where a human could not be reasonably expected to perform the calculations using the same data manually.^3^

**Definition 1.** A clinical decision support system is considered to be **AI-based** if it uses an algorithm that requires learning from some training data (e.g. Logistic regression, neural networks, etc).^4^

**Definition 2.** A clinical decision support system is considered to be **deployed** if any of the following are true:

(a) *The tool is being used within clinical practice,*
(b) *The tool has been validated by use in clinical practice, or*
(c) *The authors of the tool have made it publicly accessible*.

Hereafter we will use the acronym CDSS to refer to deployed AI clinical decision support tools.

## 2 Methods

### 2.1 Protocol and registration

This scoping review’s protocol has been published and is available on the Open Science Framework website [57].

### 2.2 Inclusion/Exclusion criteria

To be included in the scoping review, papers needed to present a deployed AI-CDSS (meeting definitions 1 and 2). Papers need to have been published in a peer reviewed journal before 31st March 2024 (with no start date) and written in English. Papers were excluded if they presented CDSSs that were clearly not related to medicine or healthcare. Tutorial, commentary, perspective, discussion and literature review papers were also excluded from the review. Where the search returned two papers that describe the same CDSS only one has been included within the review^5^. This may be due to authors publishing ‘development of’ and ‘evaluation of’ papers for the same CDSS^6^ or independent groups publishing evaluations of pre-existing CDSSs^7^. In these cases the origin paper, that is the paper that first described the CDSS, was found unless it was possible to extract all the required information from the evaluation paper. Some CDSS have been iteratively refined over time, meaning that where a more recent or updated version of an original tool was located, the older paper was rejected ^8^.

### 2.3 Search

The following bibliographic databases were searched: Pubmed, Web of Science and IEEE Xplore. The final searches were carried out in May 2025. The search terms are shown the Appendix A. PubMed was accessed using the Biopython library [29]. The PubMed and IEEE results were filtered post-search to exclude papers that were published after 1st January 2025.

### 2.4 Screening

After removing duplicates, papers were subjected to title and abstract screening against the inclusion/exclusion criteria yielding a set of papers eligible for full-text review and evaluation with respect to the inclusion/exclusion criteria. During full-text review, additional papers were identified from reviewed papers’ bibliographies (“snowballing” or citation tracing) and any papers that presented CDSSs that were also in another paper were removed and the origin paper prefered. In total, we located 130 papers for data extraction.

### 2.5 Data Extraction

Data extraction proceeded by reviewing the full-text of each paper against four key questions (the detailed criteria for each key question are given in the Appendix B):

QS0– Meta information about the paper, including year of publication, the authors’ or their institution’s geographical region, the clinical specialty of the CDSS and its intended use.

QS1– What algorithms / methods were used and what output was produced by the CDSS?

QS2– Is uncertainty considered and presented and if so, how is uncertainty presented?

QS3– How is the performance of the CDSS assessed?

Whilst not directly relevent to the presentation of uncertainty from deployed CDSSs, it is still important to consider how the performance of the models are assessed. For models that present high accuracy, people may derive reassurance that the system is reliable (by implication, certain) by looking at its summary performance. This may be misleading as there still can be significant uncertainty about a patients diagnosis, for instance because of the prevalence of the condition or because of the uncertainty associated with the creation of the model [134, 135]. This is similar to the *base rate fallacy* in traditional medical testing, where despite the fact that test can have excellent accuracy, there can still be significant uncertainty about whether or someone has a disease after a positive test [47]. We were also interested to explore how, for CDSSs that do present non-probabilistic uncertainty (for example “don’t know’ classifications), the whether performance of the uncertain outputs are explicitly assessed.

## 3 Results

### 3.1 Search Results

The search returned a total of 3897 papers across the three bibliographic databases. After removing 867 duplicates, the title and abstracts of 3831 papers were screened for inclusion. After abstract screening 306 papers were carried forward for full paper review. During the full text review 41 papers were discovered through the snow-balling process and 9 origin papers were used to extract the required data about the models. In total 411 papers where subjected to full text review, of which, 145 papers were included in this scoping review and data extraction. A PRISMA diagram is shown in figure 1. The reasons for exclusion are shown in figure 2. 138 papers were excluded as they did not present a model that met the deployed definition in definition 2. Another 60 did not meet the definition of AI (definition 1) or CDSS (definition 0). 26 papers were excluded as the model was included in a different paper. Finally, 39 papers were deemed out of scope for other reasons and 3 were not accessible to the authors.

**Figure 1:**
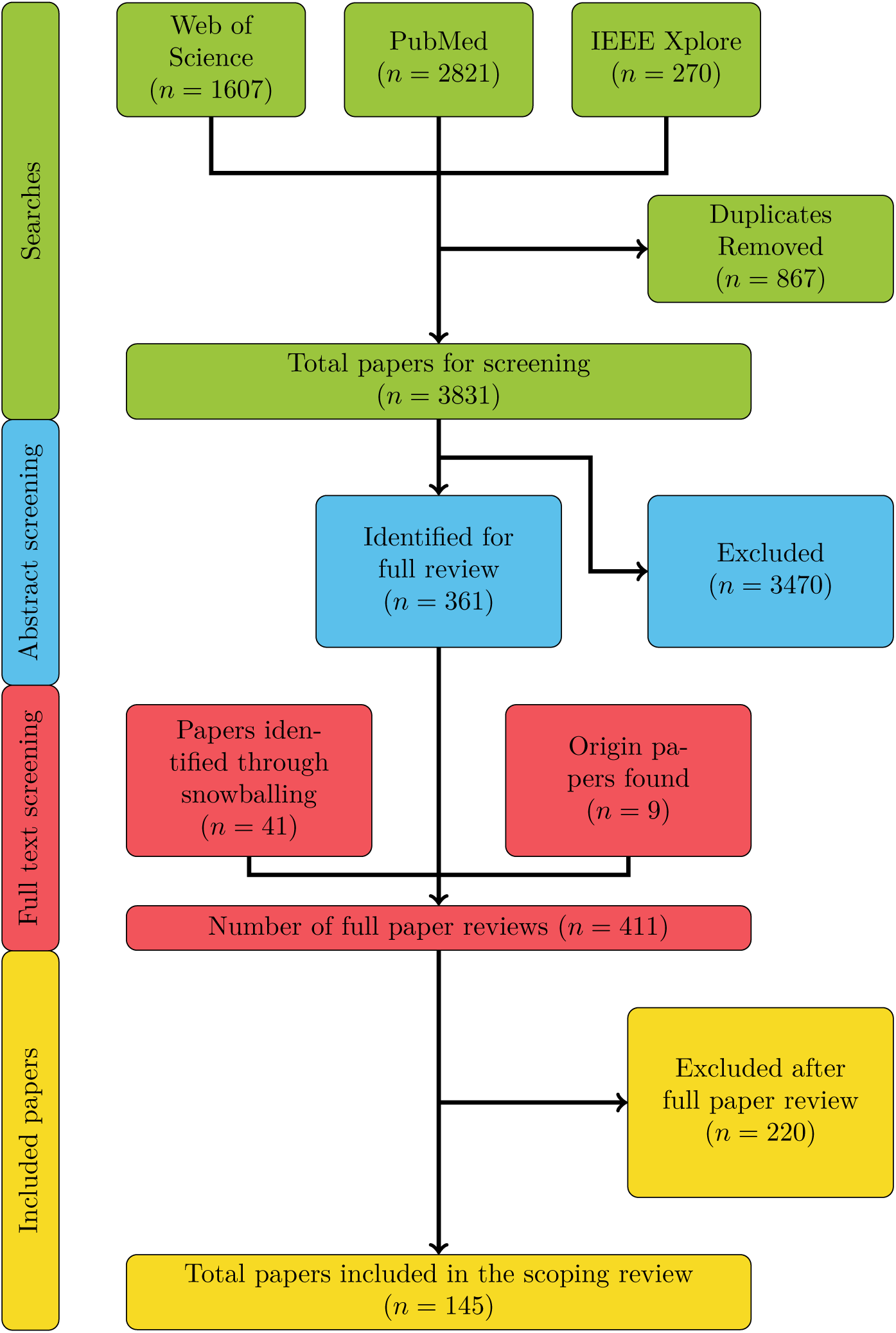
PRISMA diagram.

**Figure 2:**
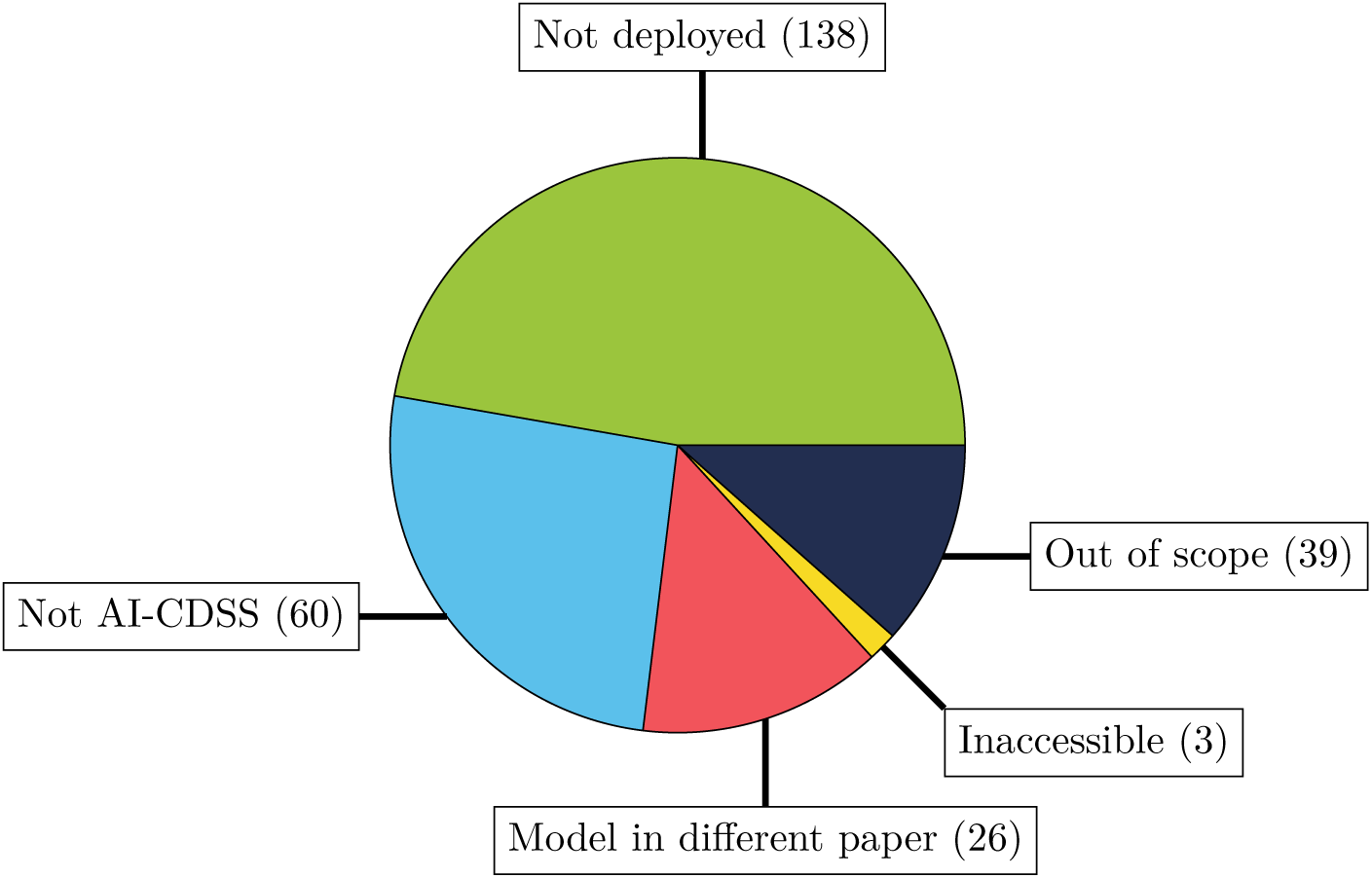
Reasons for exclusion

### 3.2 Characteristics of included studies

The number of deployed CDSSs (meeting our definition) has been increasing since the mid 2010s as shown in Figure 3, although 2024 had fewer papers than 2022 or 2023. The models found in the review covered a total of 38 different medical specialities, as shown in Figure 4. A full list of the specialities is given in Appendix C.1, Table 1. Eight of the eleven infectious disease CDSSs specifically concern the diagnosis/prognosis of COVID-19.

**Figure 3:**
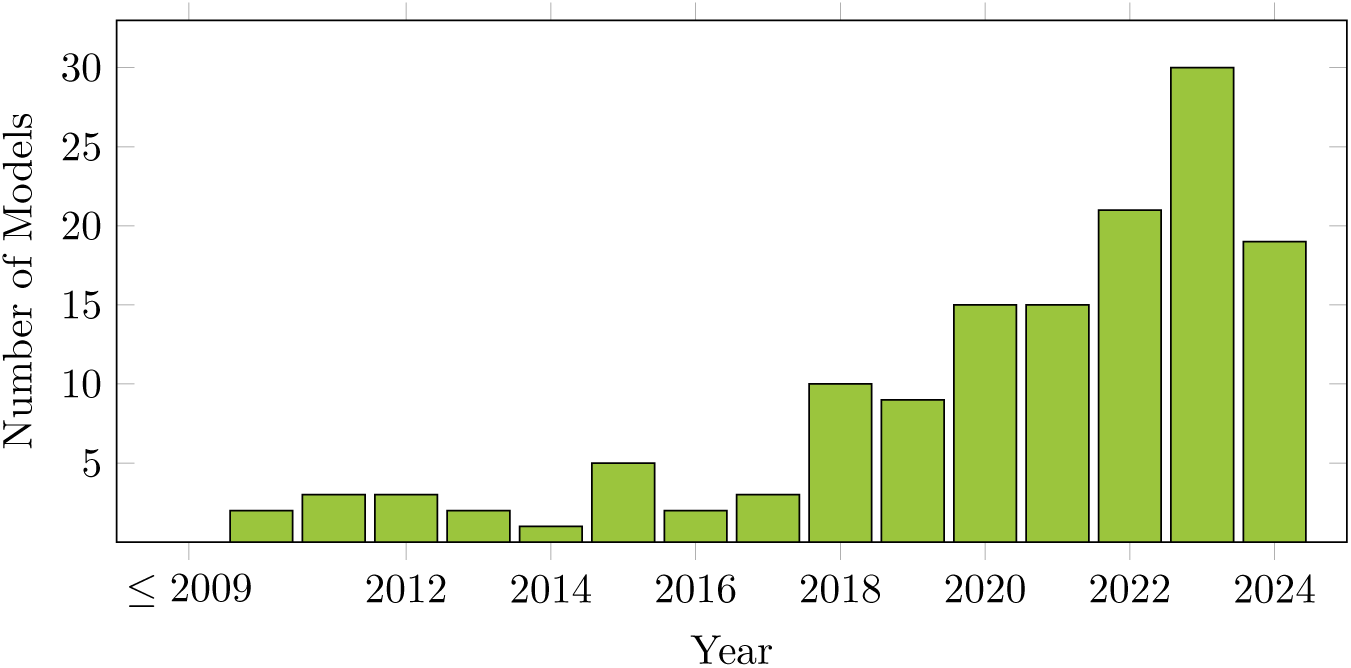
Number of models published by year. The *≤* 2009 label contains papers from all years before 2009.

**Figure 4:**
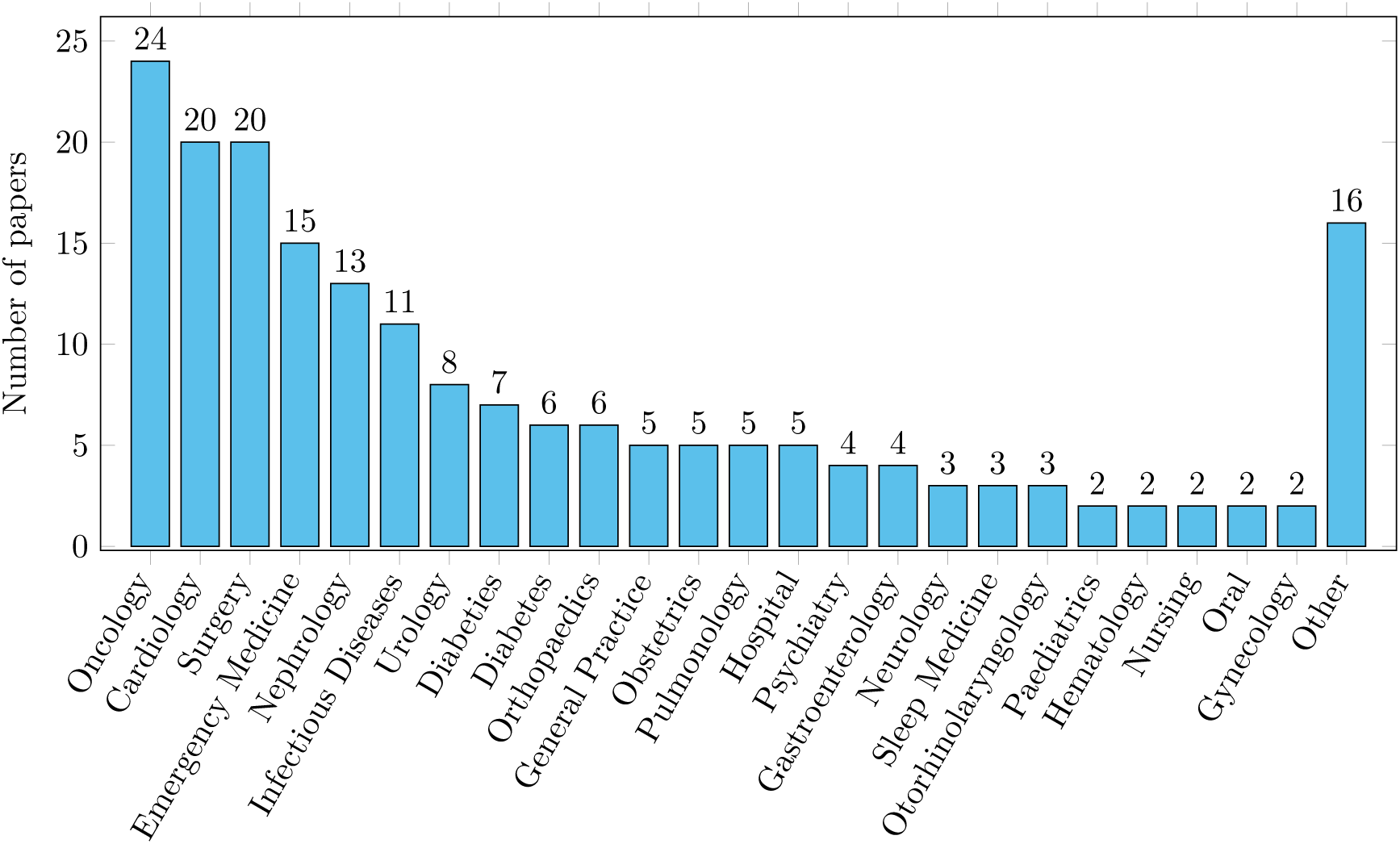
Medical specialities of the included models. The other category includes specialities for which only one model was found. A full list of specialities can be found in Table 1.

**Figure 5:**
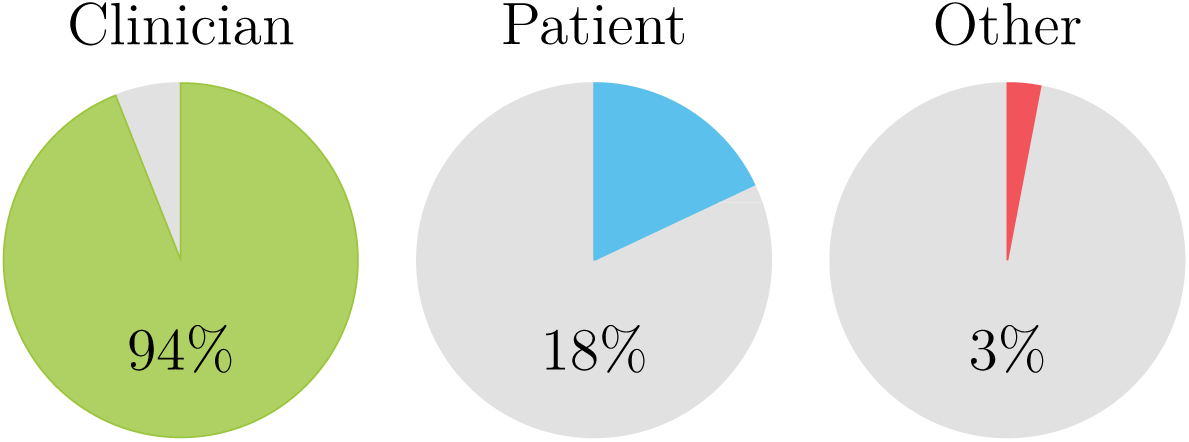
Who is the user of the CDSS

**Figure 6:** The percentage of models expected to be used by various user types.

Most of the models (136, 94%) are expected to be used by a clinician, with 28 (18%) expecting to be used by a patient with 16 expected to be used by both. 4 models had different expected users, 3 were for use by administrators [159, 161, 175] and 1 was expected to be used by a carer [141].

The purposes of the CDSSs were varied but fall into several categories, see Figure 7:

- **Prediction/diagnosis of condition** – These models predict a condition. For example, Akbulut et al. [2] predicts fetal health status and Casal-Guisande et al. [18] presents a diagnosis of sleep apnea. The output of many of these CDSSs are analogous to clinical decision making, especially in the case of diagnosis.
- **Risk of condition or clinical outcome** – These models calculate the risk of some condition occurring, sometimes the risk of the condition occurring within a specified time interval. For example, QRISK3 presents the risk of a cardio- or cerebro-vascular event occurring within a 10 year period.
- **Intervention recommendation** – These models assess the likely benefit of an intervention. For example, Lau et al. [91] present a CDSS that assesses the risk/benefit of an encephalitis vaccine and Figueiredo et al. [40] predicts the expected improvement after arthroscopic hip preservation surgery.
- **Triage/Screening** –-Assisting clinicians decide on an assessment and/or treatment service or pathway for a patient. For example, using electronic heath records [38, 42], through ‘lifestyle’ questions [188] or through medical test results [37].
- **Prediction of outcome after intervention** – A significant number of models, especially those within the surgery domain, predict the risk of adverse outcomes after a procedure or intervention is performed.
- **Monitoring/Management** – This includes papers which predict the likelihood of patients presenting to hospital emergency departments [126, 161], Umscheid, Betesh, VanZandbergen, Hanish, Tait, Mikkelsen, French, and Fuchs’s [162] sepsis early warning systems and Bertoncelli, Costantini, Persia, Bertoncelli, and D’Auria’s [12] epilepsy and seizure detection.

**Figure 7:**
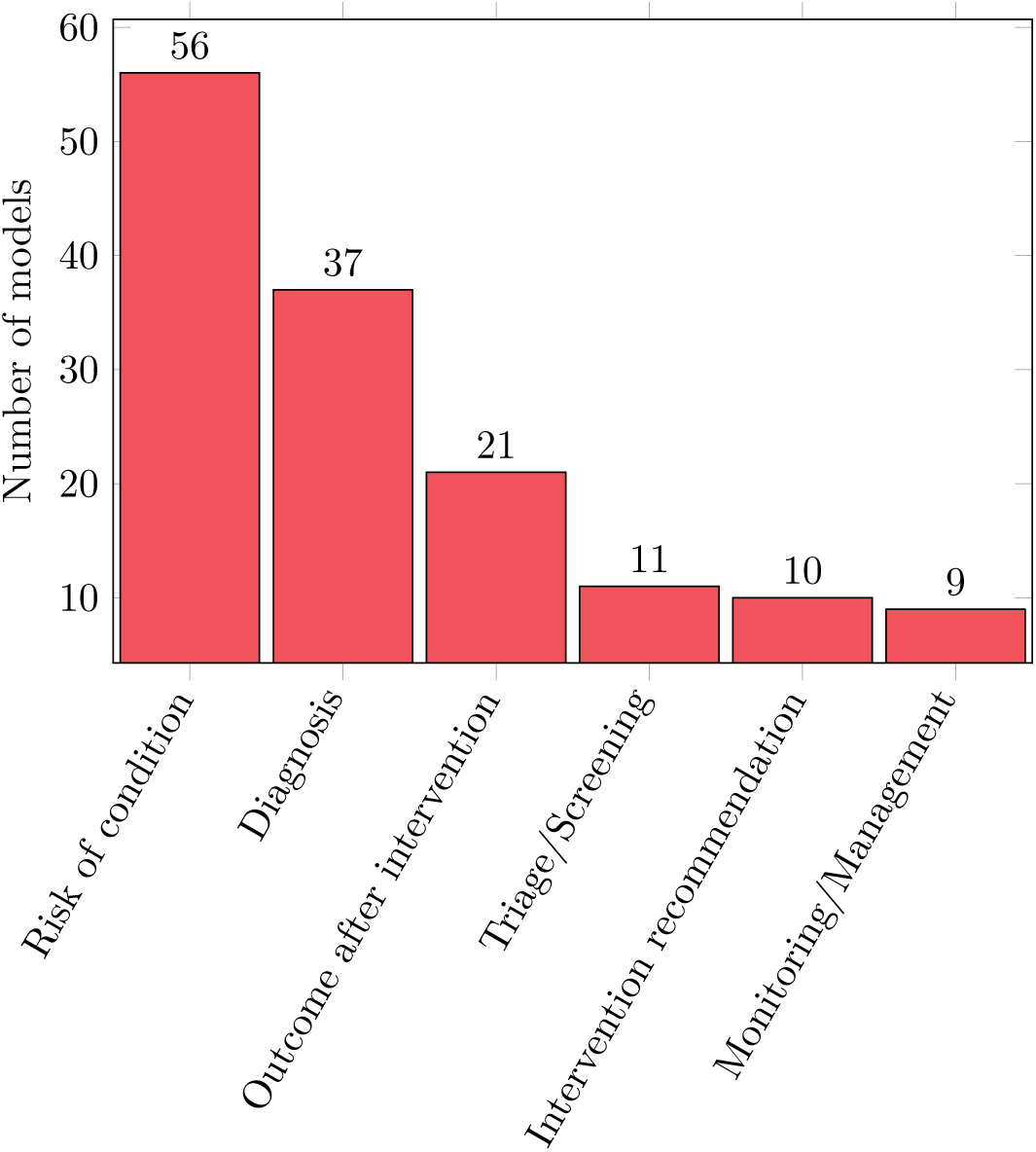
The use cases for the found CDSSs.

**Figure 8:**
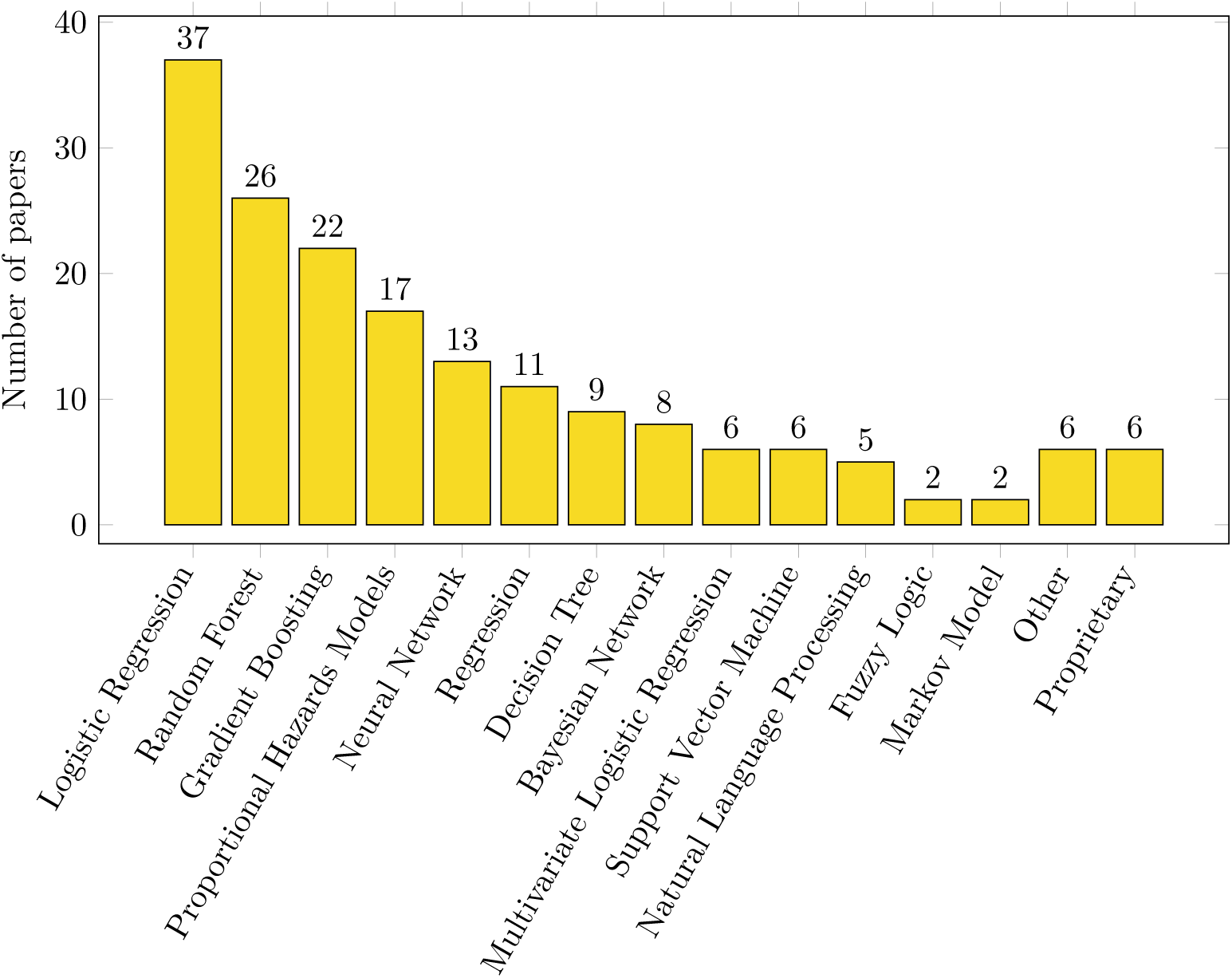
The algorithms used by the CDSSs.

Only Bertoncelli et al. presents a paper that spans multiple catagories (diagnosis and monitoring). In 23 of these papers, death is the outcome that is being predicted. The full list of papers that fall into these catagories are listed in Appendix C.2, Table 2, also listed within the same table are the death-as-outcome papers.

### 3.3 Algorithm

The most popular algorithm for deployed CDSSs was logistic regression (including related methodologies such as multivariate–, multinomial– and bayesian logistic regresion), although there was a wide range of different algorithms used. For 5 models [14, 20, 44, 46, 141] it was not possible to identify the methodology uses, this was due to the fact that the CDSSs were developed commercially and details were not described in the retrieved papers. The full list of algorithms used by the CDSSs is shown in Table 3.

### 3.4 Outputs

The majority of the CDSSs (108) present a number as the output of the model and 71 present a classification, with 33 presenting both a number and a classification. Only one CDSS presented a different output, Anand et al. [5] presented questions for a heath care professionals to ask the patients parents (since it the CDSS is for pediatric use) to help determine risk factors (example questions, include “Is [the child] in pain today” or “Does [the child] take perscription or over-the-counter medicine”). For 2 CDSSs it was not possible to find a clear description of the output delivered to the user [175, 176].

Of the 108 papers that presented a number, 90 presented a probability and of the 65 that presented a classification 57 classified the patient into risk levels (i.e. low risk/high risk). Of the classification models, only 35 papers explicitly describe the decision rule including how any classification thresholds were determined. Of these 35, 17 used an arbitrary threshold (e.g. Sun et al. [152] categorises the patient – on the basis of an event probability output – as low risk for *p <* 0.5, medium risk as 0.5 *≤ p ≤* 0.75, and high risk as *p >* 0.75). 11 papers determined a threshold by optimising various statistics: for example Huo et al. [73] optimises the decision threshold on the basis of sensitivity and specificity, Ginestra et al. [52] used positive/negative predictive value, Sher et al. [140] used Youden’s index and Patterson et al. [125] the number needed to treat. Other statistical methods were used to establish the threshold, for example, two papers [84, 90] based the threshold value of pre-existing values.

### 3.5 Uncertainty

Of the 145 papers, only 19 presented an uncertain output that wasn’t a simple probability statement of the form probability of X = 0.7 or risk of X = 70%,. These 19 included papers that presented probabilities using natural frequencies, i.e. 10 out of100 have X [13, 37, 70, 74, 141, 181]. More verbose natural language statements were also used as outputs, for example Xu et al. [181] a statement for the form *Today, in a group of 1000 people like me, 30 will have chlamydia and 970 people will not have chlamydia*. Some papers presented confidence intervals around the probability [43, 150, 167, 186] or continuous score [118, 139].

Some CDSSs presented uncertainty in a visual way; for example, Hippisley-Cox et al. [70] uses an icon array to present the probability of an event, Yu et al. [186] presents a graph of the risk level with error bars, Gardner et al. [46] used various visualisations (including icon arrays) to present risk information.

Many of these graphical outputs use colours to highlight risk levels. For example, Bilimoria et al. [15] presents their results with red for above average risk, yellow for average and green for low risk. Gonçalves et al. [55] uses colours to highlight the confidence of the output, greener denotes smaller error. Some papers present bars that grow, have a moving pointer and/or change colour [See as examples 46, 67, 148, 155, 160, 186]. Another approach used by Dihge et al. [36] and Tseng et al. [160] is to show the full distribution of the esitmated risks and highlight where the patient sits within the distribution.

### 3.6 Performance

When evaluating the performance of the CDSSs, there are two popular methods. The most popular, used by 96 CDSSs, is to use a receiver operating characteristic (ROC) curve and its associated area under the curve (AUC) metric. The second most popular method of assessing performance was to use statistics derived from a confusion matrix^10^, 66 CDSSs use this method. There were a variety of different alternatives, including: comparisons to existing methods (such as X or Y), calibration plots of expected v observed risks, Brier score, Hosmer-Lemeshow test. A full list can be found in Table 4. Many of the models use multiple different performance metrics. Figure 9 shows a plot the counts of the evaluation methods and there combinations.

**Figure 9:**
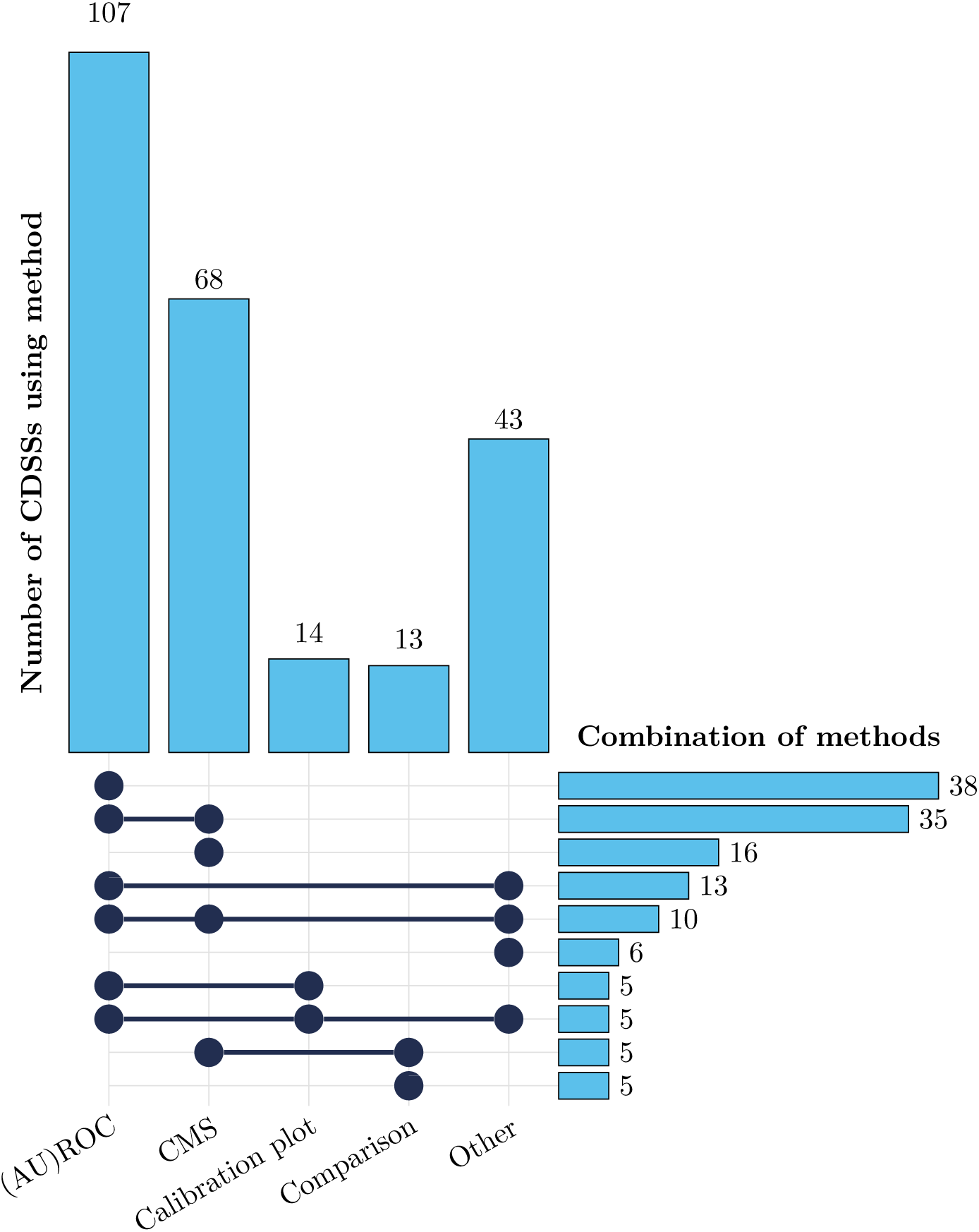
UpSet plot showing the frequency and overlap of evaluation methods used by CDSSs. The bar chart on the left shows the total number of papers using each evaluation method.. The matrix of filled circles indicates the combination of methods used with the corresponding bar on the right showing how many CDSSs used that specific combination. Only method combinations cited in more than five CDSSs are shown. The “Other” category includes evaluation methods used in fewer than 10 studies.

Of the models included in this review, only Kang et al. [82] tested how the (explicitly) outputted uncertainty, in their case a “unsure” prediction, affected the overall decision makeing process. This is acheived by checking whether uncertain predictions by the model reduced the number of false negative cases.

## 4 Discussion^11^

To frame this discussion it is useful to consider what the modal CDSS looks like and how it is may be used in practice. The tool is designed to be used by a clinician to predict the development of a health state or occurrence of a medical event in a given patient. The CDSS outputs a probability (possibly with a low/high risk classification), this probability will be presented in the form 0.74 or 74%. The performance of the CDSS will have been validated by measuring its ability to discriminate between classes using a ROC curve and through the creation of a confusion matrix to test accuracy of the binary (high/low) risk classification.

There are two distinct types of uncertainty: the aleatory uncertainty that characterises natural variability and epistemic uncertainty that covers lack of knowledge. In a medical context, aleatory uncertainty answers the question: “*How many patients with similar symptoms/histories have the disease?*”, whereas epistemic uncertainty answers *Does this patient have the disease?* When it comes to medical decision making, and for the CDSSs that present results akin to clinical reasoning, is it the latter question that needs answering. Communicating these distinct types of uncertainty via scalar probability values alone is often misunderstood, especially when describing the probability of a single patient having a disease [7, 8, 39, 50, 147, 173].

Improvements can be made by presenting information in ways people find easier to understand, such as using natural frequencies as a more intuitive way for people to understand probabilities [48, 49]. Icon arrays are another approach that can be used to communicate probabilities [45], showing natural frequency information using pictograms, however only a few models made use of these approaches despite the fact that there is a clear benefit to using them. Which approach is best is unclear, and depends on the risk, mathematical and health literacy of the user [123, 147, 168]

Another thing that is important to consider is how these probabilities should be interpreted and very often those models that present probabilities do not make it clear what exactly the probability is. For example, BASH-GN is a CDSS that assesses the risk of a patient having obstructive sleep apnea [73], that has most of the characteristics of the modal CDSS.^12^ The output of the model is a naked probability alongside a low/high-risk classification (e.g. 30.9% - low risk^13^). It is not entirely clear from the BASH-GN’s user interface what exactly this probability refers to, there are several interpretations that could be valid:

1. 31% of patients with similar symptoms suffer from sleep apnea on at least some nights,
2. 31% of nights the patient suffers from sleep apnea, or
3. 31% of the time the patient is asleep, the patient has sleep apnea episodes.

Each of these different interpretations might have a significant impact on doctors or patients decisions about further testing or treatments. The use of natural frequencies or natural language statements can help with this, for example the output could have been “*Out of 100 patients with a similar phenotype, 31 will have sleep apnea on at least some nights*” which make the output clear. However, these approaches may also be misleading as they invite a population average, whereas the key clinical decision is about what is the best decision <u>this</u> patient not what is best on average for 100 similar ones.

Many of the CDSSs present results with a high/low risk classification. As we have seen in many cases, the decision rule thresholds for classifications do not arise from a statistical methodology and most decision rules are evaluated for their performance using improper scoring rules [54]. For the modal CDSS discussed within this paper, whatever uncertain nuance is supposed to be characterised by presenting a probability with high/low risk output may be lost–especially when thresholds are set arbitrarily (i.e. without reference to clinically relevant thresholds), where thresholds have been determined from optimising classification performance (e.g. the Yourdon index) or where model-output probability calibration has not been demonstrated in advance of the imposition of a binary decision rule. A doctor might simply interpret high-risk (or a high probability) as though it implies what the correct decision should be. Such classifications are perhaps best left to epidemiologists or public health experts and not viewed as a computer science decision problem. In the framework of statistical decision theory [11, 136], the model output – a probability, or a probability distribution – should be combined with a cost/utility function for the decision to be made and the minimum-cost decision should be preferred. A difficulty with this more rigourous and formal statistical decision theoretic approach is that it is often difficult to ‘design’ or estimate a cost/utility function [63]. An exception is the use of decision curve analyses [165] where the cost of true-positive is fixed at unity and the relative cost of a false positive can be calculated (for mutually exclusive binary decisions) by deriving the net benefit over a range of threshold probabilities (model outputs).

Very few papers present uncertainty about the probabilities that they present, however this uncertainty certainly exists within all of the models. Very often such uncertainty is viewed as unhelpful at best, however it can be critical to the decision making process. BASH-GN (the CDSS predicting sleep apnea)outputting Pr = 31% implies a level of confidence that may be unwarranted. If the algorithm outputted an interval probability, the output Pr = [29, 33]% implies that the result is stable and reliable, whereas if the output was Pr = [5, 95]%, then the vacuousness of this result suggests that the algorithm should not be relied on in the decisions making process.

The most popular approaches to assessing the performance of the models are to use ROC curves or statistics derived from confusion matrices, both of these methods assess the discriminatory performance of the model. ROC curves primarily assess the trade-off between true positive rates and false positive rates across different thresholds, potentially neglecting the calibration of predicted probabilities, which is crucial for risk assessments. They also weigh errors equally and do not give information about the distribution of errors [101]. Confusion matrices, on the other hand, provide counts of true positives, true negatives, false positives, and false negatives based on a fixed threshold, which can oversimplify the performance by not capturing the uncertainty about the predictions. The important question for the user of a risk prediction model is whether an outputted probability of 31% actually implies that 31% of people have the disease. This can be done by plotting the observed vs expected risk or through a statistical means such as Hosmer-Lemeshow test or Brier score.

The use of basic binary discrimination performance metrics says nothing about clinical performance of the CDSS, especially given in most CDSSs there is significant imbalance between the classifications. For example, Cohen et al. [30] present CDSS to predict the someones suicide risk. A false negative on such a CDSS (failing to identify a suicidal individual) has much graver implications than a false positive. ROC plots cannot differentiate between the different impacts of such errors and implicitly assume that they are symmetrically consequential, whilst this is numerically convenience it is clinically nonsensical. Giving equal weighting to false positive and false negative consequences is an example of numerical convenience and clinical nonsense. Therefore, even though Cohen et al. report AUC = 0.81 (which does indicate good discriminatory performance), the tool may still be suboptimal in clinical practice if it leads to substantial and unmitigated harms.

Often overlooked when discussing uncertainty within CDSS tools is uncertainty about the model itself. Unlike the epistemic and aleatory uncertainties discussed above, this uncertainty is artefactual, it is the result of the exact dataset used, imprecision within the data, and assumptions and decisions made within the creation of the model itself. Almost all the CDSSs we investigated did not consider this as an important source of uncertainty, instead presenting a single, middle-of-the-road, model. However, there are many different CDSSs that could have been fitted from the same data [149]. Whilst some papers do optimise the data cleaning process, the selection of algorithms and hyperparameters so that the final CDSS is optimal, it should be noted that different decisions in each of these stages can lead to models that produce significantly different results [134]. Models can be highly unstable–implying large model uncertainty–especially if there is limited If the model creation process is highly unreliable, is is undesirable if for different patients a clinical decision might be made solely as an artefact of the model creation process [135]. It also needs to be acknowledged that a patient presentation is a unique instance and that their future will be influenced by a unique set of countless environmental, physiological and psychological factors in constant interplay, ergo using a single dataset to inform decision making is flawed [114, 115].

As much of the attention of AI research (and popular discourse) has moved onto the potential of large language models (LLMs) to aid in diagnosis [144]. It is critical that LLM-based CDSSs are able to correctly handle uncertainty to ensure that they do not produce factually inaccurate or harmful statements. This can can be achievied by expressing confidence about the prediction enabling users to defer to other information sources or experts when needed. This is an artefactual uncertainty, resulting from the knowledge based, training data and model parameters of the LLM, and must be treated differently to the aleatory and epistemic uncertainty discussed above. Although LLMs do have the advantage of using natural language to be able to communicate this [96]. It is also worth remembering that the ability of LLMs to pass medical exams (See Jung et al. [79] as an example), says nothing of their ability to perform these talks in the real world.

## 5 Conclusion

The promise of the increasing use of AI within medicine is that better decisions will be made sooner for (and with) patients. The downside of such an ambition is the risk that these tools are used to enable doctors to do less work whilst shifting accountability onto black-box decision making processes that purport to be evidence based. Care needs to be taken to ensure that the outputs of such CDSSs are appropriately understood, especially in a risk context. When it comes to probabilistic outputs Further research needs to be conducted to establish how best to achieve this.

Reporting protocols for AI in medicine, including TRIPOD+AI [31], CLAIM [108] and DECIDE-AI [164], do not uniformly consider either the uncertainty associated with the outputs of a model nor how this is presented in a CDSS. This is bacause most of the protocols focus upon model training/development and testing/validation and not on the deployment. The informatics of end-to-end CDSS development and deployed optimisation is a more complex problem involving risk/uncertainty communication, decision theory and human-computer interaction considerations.

The development of CDSSs needs to not be seen as a tournament of algorithms competing to be the most empirically correct, but as a part of system to improve the medical decision making process in general. Careful communication of risk and uncertainty is a key component of that process.

## Data Availability

n/a

## A Search terms

### A.1 PubMed Search Term

“ML”[Title/Abstract] OR “Artificial intelligence”[Title/Abstract] OR “AI”[Title/Abstract] OR “machine intelligence”[Title/Abstract] OR “machine learning”[Title/Abstract] OR “intelligent *”[Title/Abstract] OR “expert system”[Title/Abstract] OR “neural network”[Title/Abstract] OR “natural language processing”[Title/Abstract] OR “generative AI”[Title/Abstract] OR “deep learning”[Title/Abstract] OR “bayesian”[Title/Abstract] OR “fuzzy logic”[Title/Abstract] OR “Artificial Intelligence”[MeSH Terms] AND (“predictive modelling”[Title/Abstract] OR “prediction modelling”[Title/Abstract] OR “prognostic modelling”[Title/Abstract] OR “Decision Support Tool”[Title/Abstract] OR “decision support system”[Title/Abstract] OR “risk prediction”[Title/Abstract] OR “decision support systems, clinical”[MeSH Terms]) AND (“Journal Article”[Publication Type]) AND (“uncertainty”[Title/Abstract] OR “risk”[Title/Abstract]) AND (“English”[Language]) NOT “imaging”[Title/Abstract] NOT “image”[Title/Abstract] NOT “vision”[Title/Abstract] NOT “literature review”[Title/Abstract] NOT “scoping review”[Title/Abstract] NOT “systematic review”[Title/Abstract] NOT “environment*”[Title/Abstract] NOT “veterinary”[Title/Abstract] NOT “Organisims”[Title/Abstract] NOT “Drug”[Title/Abstract]

### A.2 Web of Science search term

The search can be accessed at: https://www.webofscience.com/wos/woscc/summary/056d9f82-4755-4801-b331-50c6 relevance/1

#### A.3 IEEE Xplore Search Term

((“ML” OR “Artificial intelligence” OR “AI” OR “machine intelligence” OR “machine learning” OR “expert system” OR “neural network” OR “natural language processing” OR “generative AI” OR “deep learning” OR “bayesian” OR “fuzzy logic” OR “Artificial Intelligence”) AND (“predictive modelling” OR “prediction modelling” OR “prognostic modelling” OR “Decision Support Tool” OR “decision support system” OR “risk prediction”) AND (“uncertainty” OR “risk”) NOT (“imaging” OR “image” OR “vision” OR “literature review” OR “scoping review” OR “systematic review” OR “environment*” OR “veterinary” OR “Organisms” OR “Drug”))

## B Questions

The use of square brackets indicates that the answer was expected to be a categorical answer. In all cases *Other* answers are followed up by establishing what the other is.

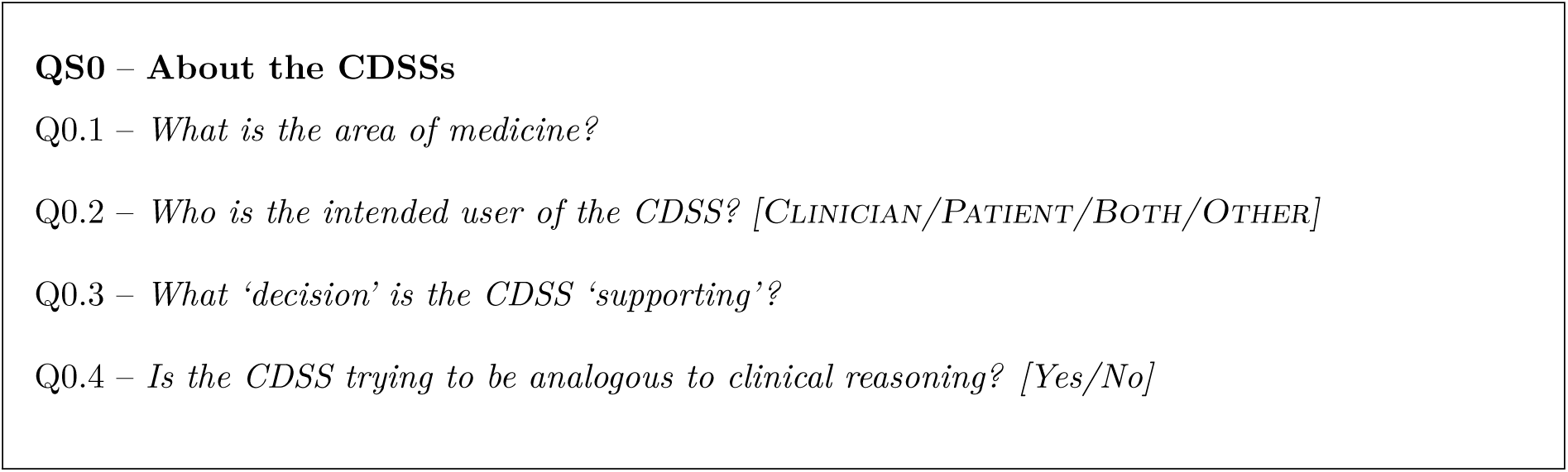

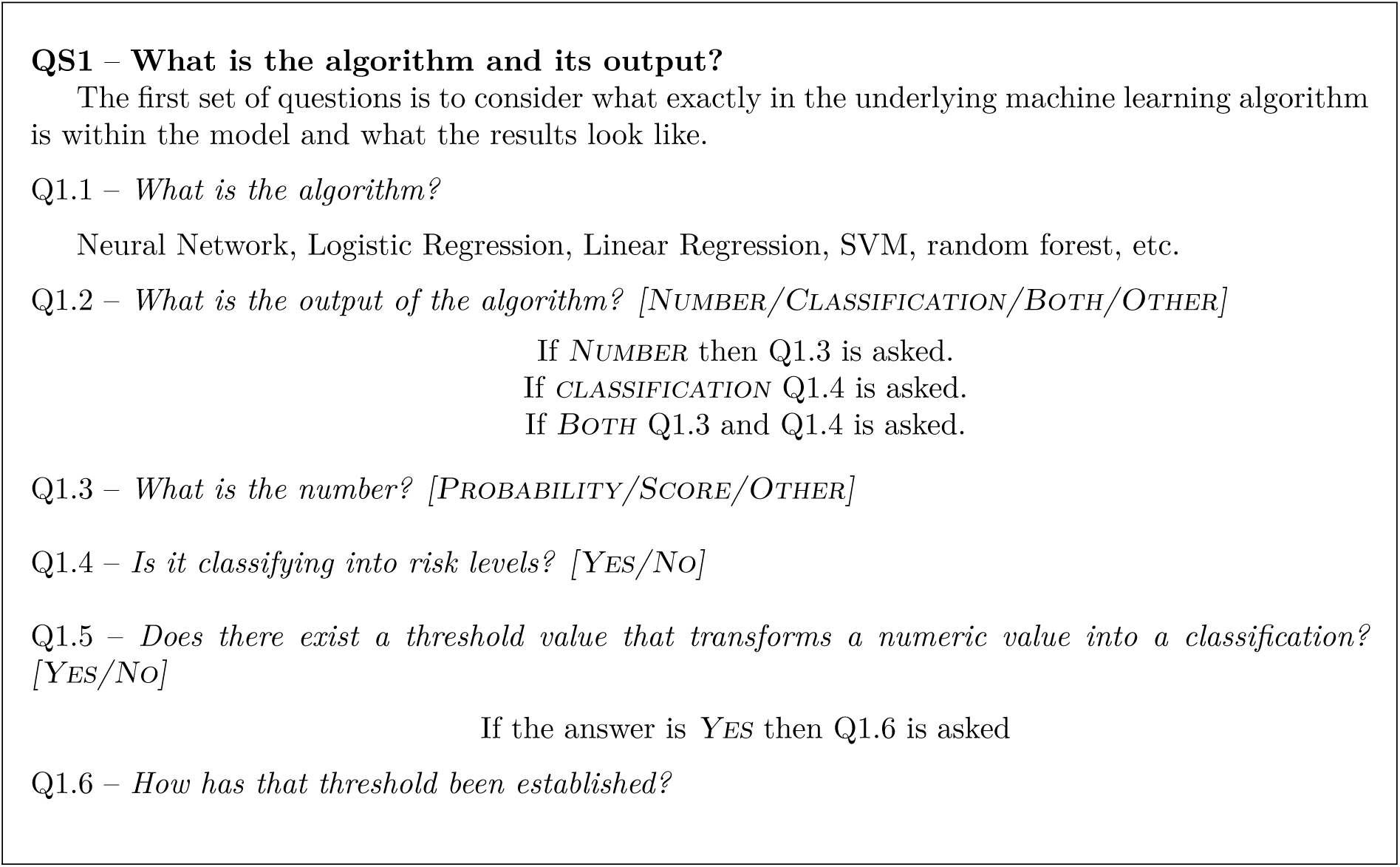

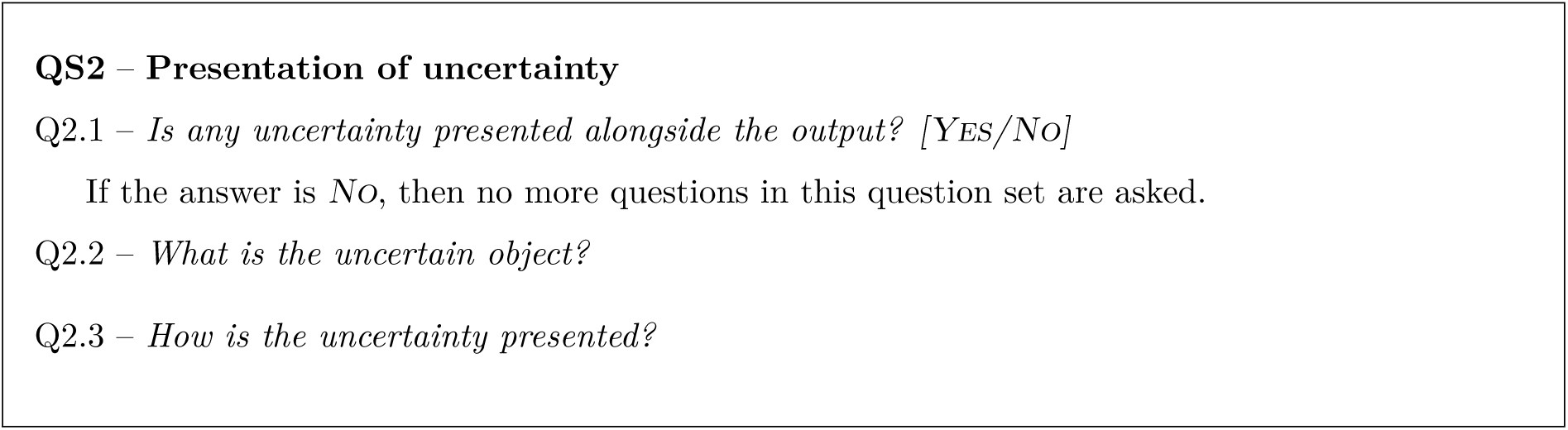

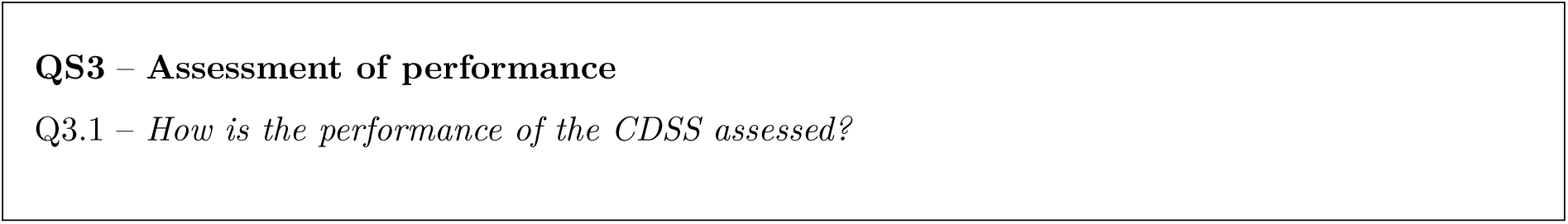

## C Data Tables

### C.1 Medical specialities

**Table 1:** Medical Specialities of the included models.

| Specialty | Count | Reference | Specialty | Count | Reference |
| --- | --- | --- | --- | --- | --- |
| Oncology | 24 | 3, 9, 27, 36, 43, 44, 55, 83, 86, 95, 97, 105, 110, 124, 140, 142, 150, 151, 160, 166, 172, 184, 185, 188 | Oral | 2 | 33, 160 |
| Cardiology | 20 | 22, 38, 41, 60, 68, 70, 78, 84, 102, 111, 113, 120, 130, 131, 148, 159, 163, 170, 174, 186 | Paediatrics | 2 | 5, 182 |
| Surgery | 20 | 1, 13, 14, 15, 17, 55, 61, 64, 65, 92, 94, 107, 113, 116, 120, 127, 128, 138, 167, 171, 175 | Hematology | 2 | 78, 131 |
| Emergency Medicine | 15 | 13, 16, 35, 56, 74, 106, 126, 127, 128, 139, 153, 155, 161, 167, 176 | Neonatology | 1 | 98 |
| Nephrology | 13 | 10, 28, 66, 67, 81, 89, 99, 117, 119, 143, 156, 157, 169 | Malnutrition | 1 | 158 |
| Infectious Diseases | 11 | 4, 23, 34, 40, 52, 76, 77, 146, 162, 179, 184 | Sports Medicine | 1 | 88 |
| Urology | 8 | 9, 12, 24, 40, 74, 81, 119, 143, 150, 151, 166 | Orthopedics | 1 | 88 |
| Diabetics | 7 | 37, 71, 104, 137, 141, 145, 171 | Outpatients | 1 | 155 |
| Diabetes | 6 | 26, 40, 46, 122, 180, 187 | Geriatric Care | 1 | 161 |
| Orthopaedics | 6 | 21, 64, 65, 93, 121, 129 | Rheumatology | 1 | 90 |
| Hospital | 5 | 85, 118, 125, 152, 177 | Tropical Medicine | 1 | 56 |
| General Practice | 5 | 42, 46, 106, 126, 153 | Neurosurgery | 1 | 138 |
| Obstetrics | 5 | 2, 37, 100, 178, 180 | Sexual Health | 1 | 181 |
| Pulmonology | 5 | 20, 53, 78, 94, 131 | Psychology | 1 | 105 |
| Psychiatry | 4 | 30, 74, 109, 132 | Transplant | 1 | 51 |
| Gastroenterology | 4 | 58, 61, 103, 183 | Hepatology | 1 | 51 |
| Sleep Medicine | 3 | 59, 73, 82 | Travel Medicine | 1 | 91 |
| Neurology | 3 | 12, 24, 74 | Social Care | 1 | 106 |
| Otorhinolaryngology | 3 | 18, 59, 73 | Palliative Care | 1 | 124 |

### C.2 Use case of the CDSSs

**Table 2:**
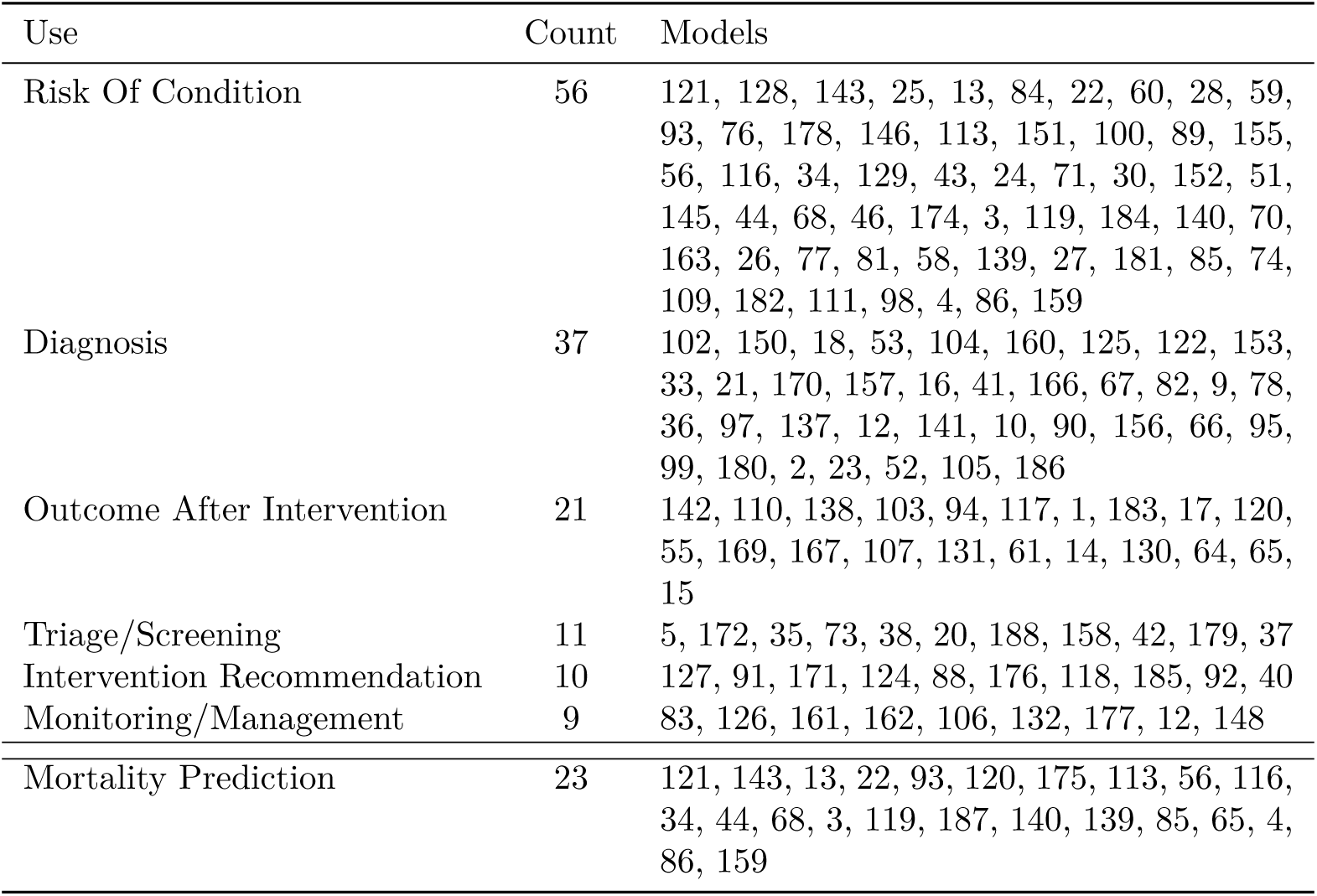
The use cases of the CDSSs.

| Use | Count | Models |
| --- | --- | --- |
| Risk Of Condition | 56 | 121, 128, 143, 25, 13, 84, 22, 60, 28, 59, 93, 76, 178, 146, 113, 151, 100, 89, 155, 56, 116, 34, 129, 43, 24, 71, 30, 152, 51, 145, 44, 68, 46, 174, 3, 119, 184, 140, 70, 163, 26, 77, 81, 58, 139, 27, 181, 85, 74, 109, 182, 111, 98, 4, 86, 159 |
| Diagnosis | 37 | 102, 150, 18, 53, 104, 160, 125, 122, 153, 33, 21, 170, 157, 16, 41, 166, 67, 82, 9, 78, 36, 97, 137, 12, 141, 10, 90, 156, 66, 95, 99, 180, 2, 23, 52, 105, 186 |
| Outcome After Intervention | 21 | 142, 110, 138, 103, 94, 117, 1, 183, 17, 120, 55, 169, 167, 107, 131, 61, 14, 130, 64, 65, 15 |
| Triage/Screening | 11 | 5, 172, 35, 73, 38, 20, 188, 158, 42, 179, 37 |
| Intervention Recommendation | 10 | 127, 91, 171, 124, 88, 176, 118, 185, 92, 40 |
| Monitoring/Management | 9 | 83, 126, 161, 162, 106, 132, 177, 12, 148 |
| Mortality Prediction | 23 | 121, 143, 13, 22, 93, 120, 175, 113, 56, 116, 34, 44, 68, 3, 119, 187, 140, 139, 85, 65, 4, 86, 159 |

### C.3 Algorithms used by the CDSSs

**Table 3:** Algorithm used by CDSSs.

| Algorithm | Count | Paper Reference |
| --- | --- | --- |
| Logistic Regression | 37 | 9, 12, 16, 17, 21, 33, 34, 40, 41, 43, 55, 56, 67, 71, 73, 76, 88, 89, 100, 107, 113, 116, 118, 120, 126, 129, 130, 131, 146, 151, 153, 155, 157, 161, 162, 166, 167, 170, 169, 176, 175, 183 |
| Random Forest | 26 | 2, 23, 27, 42, 52, 58, 66, 67, 74, 77, 81, 85, 86, 92, 95, 99, 105, 109, 138, 139, 158, 159, 179, 180, 181, 182 |
| Gradient Boosting | 22 | 1, 22, 28, 53, 59, 60, 76, 83, 86, 92, 93, 94, 103, 104, 117, 122, 124, 125, 138, 159, 160, 171 |
| Proportional Hazards Models | 17 | 3, 10, 26, 66, 70, 90, 119, 130, 140, 156, 162, 163, 174, 184, 185, 187, 188 |
| Neural Network | 13 | 12, 36, 51, 61, 67, 97, 109, 132, 137, 145, 152, 159, 177 |
| Regression | 11 | 4, 9, 12, 15, 16, 17, 21, 33, 34, 40, 41, 43, 55, 56, 64, 65, 67, 71, 73, 76, 88, 89, 98, 100, 107, 111, 113, 116, 118, 120, 126, 129, 130, 131, 146, 151, 153, 155, 157, 159, 161, 162, 166, 167, 170, 169, 176, 175, 182, 183, 186 |
| Decision Tree | 9 | 13, 18, 83, 84, 106, 110, 142, 150, 176 |
| Bayesian Network | 8 | 25, 91, 102, 121, 127, 128, 143, 172 |
| Multivariate Logistic Regression | 6 | 9, 71, 118, 130, 131, 162 |
| Support Vector Machine | 6 | 30, 37, 67, 86, 109, 159 |
| Private | 6 | 14, 20, 44, 46, 68, 141 |
| Natural Language Processing | 5 | 30, 38, 78, 83, 106 |
| Markov Model | 2 | 24, 82 |
| Fuzzy Logic | 2 | 18, 35 |
| Voting Classifier | 1 | 159 |
| Principle Component Analysis | 1 | 12 |
| Learning From Examples Using Rough Sets (Lers) | 1 | 178 |
| Arden Syntax Medical Logic Modules | 1 | 5 |
| Ensemble Prediction Model | 1 | 138 |
| Reinforcement Learning | 1 | 148 |

### C.4 Evaluation metrics used by the CDSSs

**Table 4:** Metrics used to assess the performance of the CDSSs.

| Metric | Count | Models |
| --- | --- | --- |
| Calibration Plot | 15 | 15, 43, 58, 60, 66, 88, 93, 94, 100, 120, 140, 151, 159, 185, 188 |
| Comparison To Existing Practice | 14 | 1, 20, 25, 35, 38, 41, 52, 68, 83, 91, 106, 124, 162, 166 |
| Brier Score | 8 | 4, 15, 60, 127, 128, 151, 159, 160 |
| Hosmer-Lemeshow | 7 | 15, 71, 113, 127, 128, 151, 159 |
| User Views | 2 | 141, 148 |
| Hazard Ratio | 1 | 122 |
| Mann-Whitney U Test | 1 | 88 |
| Wilcoxon Rank-Sum Test | 1 | 9 |
| Unclear | 1 | 46 |
| D-Statistic | 1 | 70 |
| R2 Test | 1 | 70 |
| (AU)ROC | 108 | 3, 4, 9, 10, 14, 15, 16, 17, 18, 22, 23, 25, 27, 30, 33, 34, 36, 37, 40, 44, 51, 53, 55, 56, 58, 59, 60, 61, 64, 65, 66, 67, 70, 71, 73, 74, 76, 77, 78, 81, 83, 85, 86, 88, 90, 92, 93, 95, 99, 97, 98, 102, 103, 104, 105, 107, 111, 113, 116, 117, 118, 120, 119, 121, 125, 127, 128, 129, 130, 131, 132, 137, 138, 139, 140, 142, 143, 145, 146, 150, 151, 152, 153, 155, 157, 158, 159, 160, 161, 163, 167, 172, 171, 170, 169, 174, 176, 175, 179, 180, 181, 182, 183, 184, 185, 186, 187, 188 |
| (AU)PRC | 7 | 59, 85, 92, 104, 158, 163, 180 |
| CMS | 68 | 1, 2, 4, 5, 12, 13, 14, 16, 18, 20, 21, 23, 24, 25, 27, 28, 30, 35, 38, 42, 44, 51, 52, 53, 55, 56, 59, 73, 74, 76, 78, 82, 84, 85, 89, 90, 92, 93, 97, 102, 104, 105, 109, 110, 116, 117, 125, 126, 131, 138, 139, 143, 150, 155, 157, 160, 161, 163, 172, 171, 170, 169, 176, 177, 178, 179, 181, 182 |
| NNT | 1 | 125 |
| DCA | 6 | 9, 33, 51, 175, 185, 186 |
| $\chi^2$ Test | 5 | |
| 26 68 97 124 156 |  |  |

## Footnotes

1 www.qrisk.org/

2 These are of the style: “In other words, in a crowd of 100 people with the same risk factors as you, *N* are likely to have a heart attack or stroke within the next 10 years.”

3 For example, a computerised version of the PHQ9 questionnaire [87] that simply reports the total score for screening would be excluded since it could be calculated manually and on it’s own, a computer-based implementation of PHQ9 is not prognostic.

4 As opposed to an algorithm that been derived from pre-existing logical rules.

5 Very often this would be the development paper even though it itself might not be in scope as it fails to meet definition 2

6 Jauk et al. present two papers of their delirium prediction model, in [74] the model is introduced whereas Jauk et al. 75 qualitative explores the use if the algorithm within a clinical setting. As such, only [74] is included within this review.

7 For example, [6] independently assess the performance of EuroScore II in patients with structural deterioration of aortic bioprostheses.

8 For example, EuroSCORE II [113] was included over the original EuroSCORE model [112]

9 For this study, logistic regression includes related methodologies such as multivariate–, multinomial– and bayesian logistic regresion.

10 Such as accuracy, sensitivity/specificity, precision/recall, F1 score etc

11 The use of example papers within this section is in no way intended to denigrate particular authors or papers nor to be reflexive of the merits of the CDSS presented within them.

12 It is accessible at https://c2ship.org/bash-gn-metric/

13 This result is produced with the following inputs: female, aged 54, neck circumference 24cm, weight 90kg, height 1.56m, with high blood pressure and snoring as loud as talking.

